# Forecasting the Wuhan coronavirus (2019-nCoV) epidemics using a simple (simplistic) model

**DOI:** 10.1101/2020.02.04.20020461

**Authors:** Slav W. Hermanowicz

## Abstract

Confirmed infection cases in mainland China were analyzed using the data up to January 28, 2020 (first **13 days** of reliable confirmed cases). For the first period the cumulative number of cases followed an exponential function. However, from January 28, we discerned a downward deviation from the exponential growth. This slower-than-exponential growth was also confirmed by a steady decline of the effective reproduction number. A backtrend analysis suggested the original basic reproduction number *R*_0_ to be about 2.4 to 2.5. As data become available, we subsequently analyzed them during three consecutive periods obtaining a sequence of model predictions. All available data up were processed the same way. We used a simple logistic growth model that fitted very well with ***all* data**. Using this model and the three sets of data, we estimated **maximum cases** as about 21,000, 28,000 and 35,000 cases refining these predictions in near-real time. With slightly different approach (linearization in time) the estimate of maximum cases was even higher (about 65,000). Although the estimates of maximum cases increase as more data were reported all models show reaching a peak in **mid-February** in contrast to the unconfined exponential growth. These predictions do not account for any possible other secondary sources of infection.

## Introduction

Recent outbreak of a novel coronavirus (designated 2019-nCoV) originated in Wuhan, China raised serious public health concerns and many human tragedies. To manage the epidemics resulting from virus spreading across China and other countries forecasting the occurrence of future cases is extremely important. Such forecasting is very complicated and uncertain since many factors are poorly understood or estimated with a large possible error. Two major factors include spatial movement of virus carriers (i.e., infected individuals) and the basic reproduction number *R*_0_ (average number of new infections originated from an infected individual).

As a result of great public attention to the virus spreading several approaches were recently reported attempting to model the epidemics and infection dynamics (Chen et al., 2020; Hui et al., 2020; Imai, Cori, et al., 2020; Imai, Dorigatti, Cori, Donnelly, et al., 2020; Li et al., 2020; Linton et al., 2020; Liu et al., 2020; Shen, Peng, Xiao, & Zhang, 2020; Wu, Leung, & Leung, 2020; Zhang & Wang, 2020b; Zhao et al., 2020a). These models are certainly useful to understand the implications of various quarantine procedures, public health actions, and possible virus modifications. However, the reported models are, by its nature, very complicated with numerous assumptions and requiring many parameters values of which are not known with good accuracy. As a result, some predictions (Nishiura et al., 2020) were more educated guesses although eventually they were confirmed qualitatively.

In this work, we present the results of fitting a very simple (perhaps even simplistic) model to the available data and a forecast of new infections.

### Logistic Model and its Application

The logistic model has been used in population dynamics and specifically in epidemics for a long time (Bailey, 1950; Bangert, Molyneux, Lindsay, Fitzpatrick, & Engels, 2017; Cockburn, 1960; Jowett, Browning, & Haning, 1974; Koopman, 2004; Mansfield & Hensley, 1960; Waggoner & Aylor, 2000). Mathematically, the model describes dynamic evolution of a population *P* (in our case the number of infected individuals) being controlled by the growth rate *r* and population capacity *K* due to limiting resources. In continuous time *t* the change of *P* is

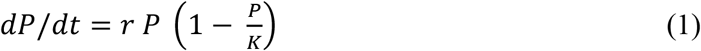

Initially, the growth of *P* is close to exponential since the term (1 − *P/K*) is almost one. When *P* becomes larger (commensurate with *K*) the growth rate slows down with

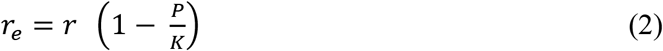

becoming an effective instantaneous growth rate.

In discrete time, more appropriate to daily reported infection cases, the logistic model becomes

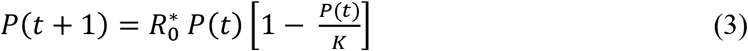

where *P(t)* and *P(t+1)* are populations on consecutive days, *R* _0_^*^ is the growth rate (basic reproduction number in epidemiology) at the beginning of the logistic growth, and *K* is the limiting population. When plotted as a function of time *t*, both Eqs. (1) and (3) result in a classic sigmoidal curve with

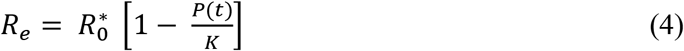

being the effective reproduction rate at time *t*.

The logistic model may be adequate for the analysis of mainland China since the country can be treated as a unit where a vast majority of cases occurred without any ***significant*** “import” or “export” of cases.

### Data Analysis

We analyzed infection cases in mainland China as reported by the National Health Commission of the People’s Republic of China (www.nhc.gov.cn/xcs/xxgzbd/gzbd_index.shtml). As of the time of writing the number of cases is shown in Table 1. The data are also plotted in Figure *1*. The first column in Table 1 represents days from the beginning of the outbreak. There is a considerable controversy as to the exact date of the outbreak with most reports pointing to mid-December (Li et al., 2020; Wang, Horby, Hayden, & Gao, 2020) while one analysis suggest multiple sources of original infection (Nishiura et al., 2020). Initially, the outbreak was not recognized and number of ***confirmed*** cases is not fully known (P. Wu et al., 2020). Initially, (Figure *1*), the number of cases increased exponentially. This feature was also clearly recognized in a previous report (Zhao et al., 2020b).

**Figure 1.**
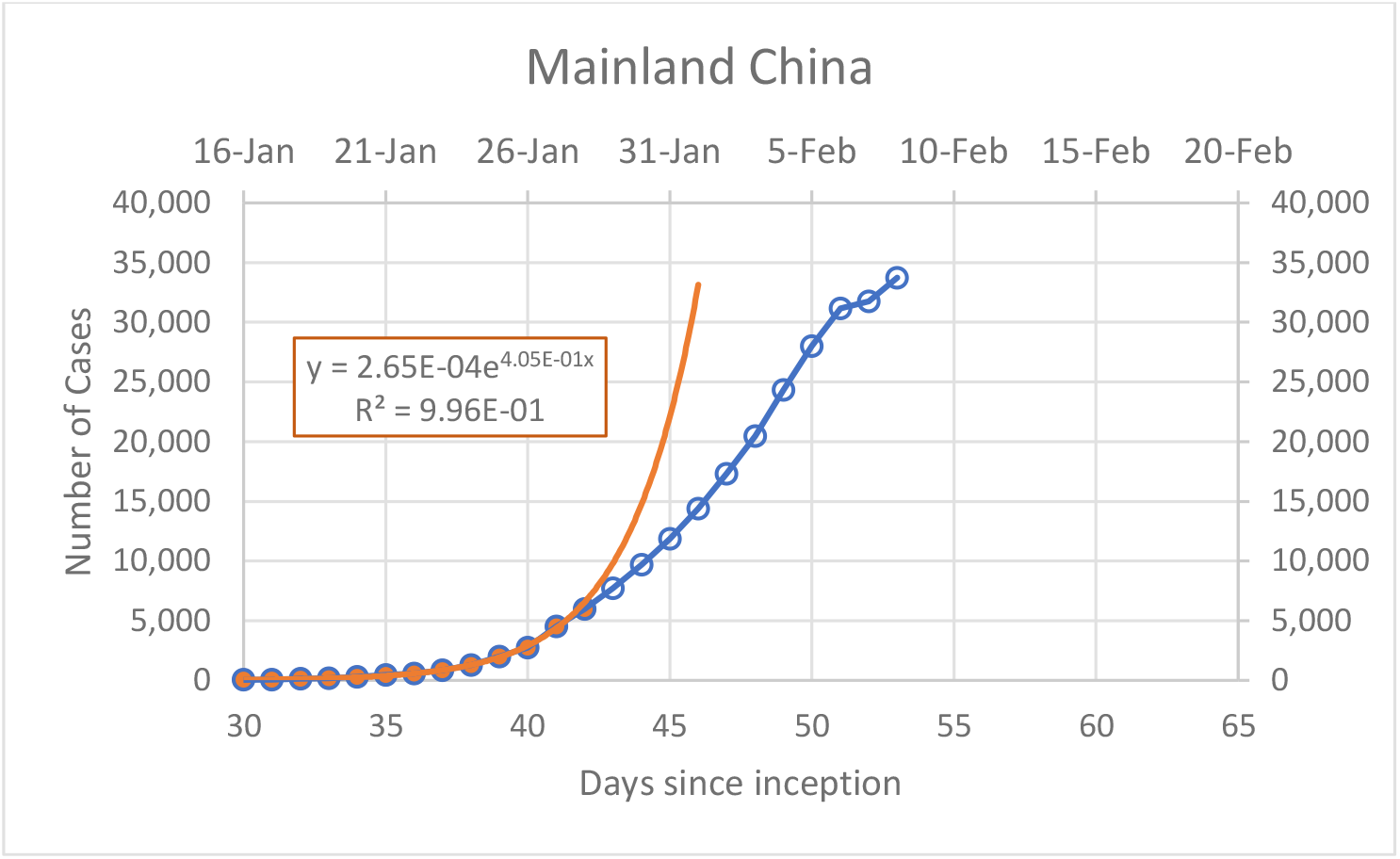
Evolution of recorded infection cases. From day 42 there is a significant slowing of the infections.

**Table 1.** Reported cases in mainland China

| Day | Date | Cases | $R_e$ |
| --- | --- | --- | --- |
| 30 | January 16, 2020 | 45 |  |
| 31 | January 17, 2020 | 62 | 1.38 |
| 32 | January 18, 2020 | 121 | 1.95 |
| 33 | January 19, 2020 | 198 | 1.64 |
| 34 | January 20, 2020 | 291 | 1.47 |
| 35 | January 21, 2020 | 440 | 1.51 |
| 36 | January 22, 2020 | 571 | 1.30 |
| 37 | January 23, 2020 | 830 | 1.45 |
| 38 | January 24, 2020 | 1,287 | 1.55 |
| 39 | January 25, 2020 | 1,975 | 1.53 |
| 40 | January 26, 2020 | 2,744 | 1.39 |
| 41 | January 27, 2020 | 4,515 | 1.65 |
| 42 | January 28, 2020 | 5,974 | 1.32 |
| 43 | January 29, 2020 | 7,711 | 1.29 |
| 44 | January 30, 2020 | 9,692 | 1.26 |
| 45 | January 31, 2020<sup>(1)</sup> | 11,860 | 1.22 |
| 46 | February 1, 2020 | 14,380 | 1.21 |
| 47 | February 2, 2020 | 17,307 | 1.20 |
| 48 | February 3, 2020<sup>(2)</sup> | 20,467 | 1.18 |
| 49 | February 4, 2020 | 24,324 | 1.19 |
| 50 | February 5, 2020 | 28,018 | 1.15 |
| 51 | February 6, 2020<sup>(3)</sup> | 31,161 | 1.11 |
| 52 | February 7, 2020 | 31,774 | 1.02 |
| 53 | February 8, 2020 | 33,738 | 1.06 |
| 54 | February 9, 2020 |  |  |

(1), (2), (3) – ends of Periods 1, 2, and 3. Data beyond Feb. 6 were not used in model fitting and predictions

However, starting from January 28 (day 42) there was a marked deviation from the exponential curve, in line with logistic growth. This decline of the ***growth rate*** is clearly demonstrated by a steady decline of the effective reproduction number *R*_*e*_ (fourth column in Table 1) calculated simply as a ratio of cases on consecutive days

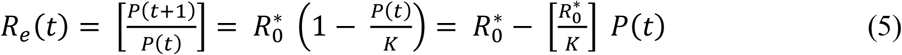

Eq. (5) also shows a linearization of the discrete logistic model (Eq. (3)) that was used to estimate the *R*_*e*_ *(t=30) = R*^*0\**^ and capacity *K*. An example of such linearization for Period 1 is shown in Figure *2* with the initial 16 points (blue points and line in Figure *2*). The same figure also shows linearization using for Period 2 (orange points and line in Figure *2*). For all periods the effective reproduction numbers decrease in time.

**Figure 2.**
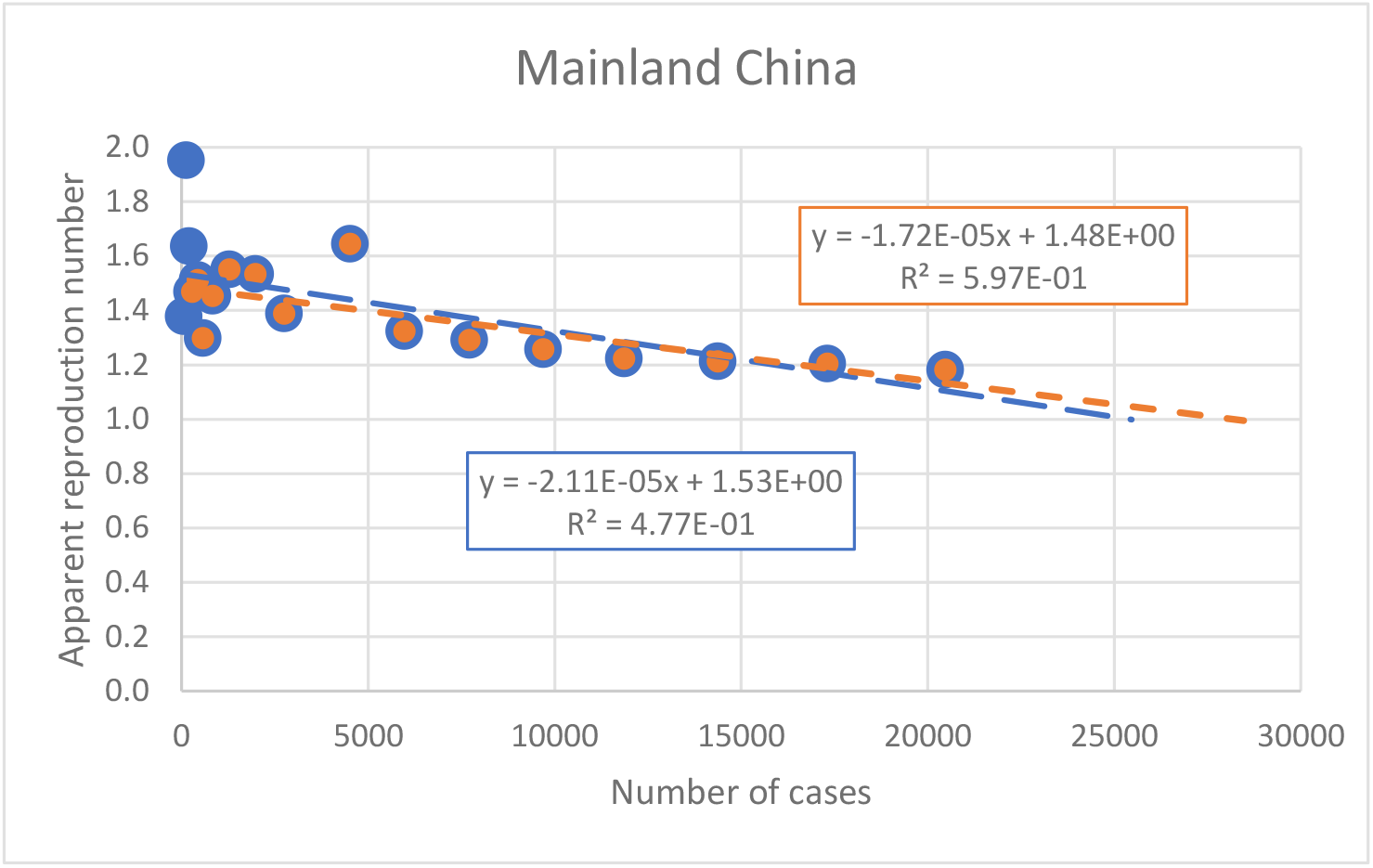
Linearization of the effective reproduction number with number of cases following Eq. (5) for Periods 1 and 2

As new data were reported daily, we followed with subsequent analysis in near-real time with three periods as shown in **Table *2***

In contrast, a simple linearization of *R*_*e*_ in time (Figure *3*) back-estimated the original basic reproduction number *R*_0_ at about 2.4 to 2.5, agreeing well with other values recently reported (Imai, Dorigatti, Cori, Riley, & Ferguson, 2020; Liu et al., 2020; Majumder & Mandl, 2020; Zhang & Wang, 2020a, 2020b; Zhao et al., 2020a). The discrepancy between these two estimates can be attributed a potential loss of virulence of the virus but most likely due to extreme measures to contain virus spread in China.

**Figure 3.**
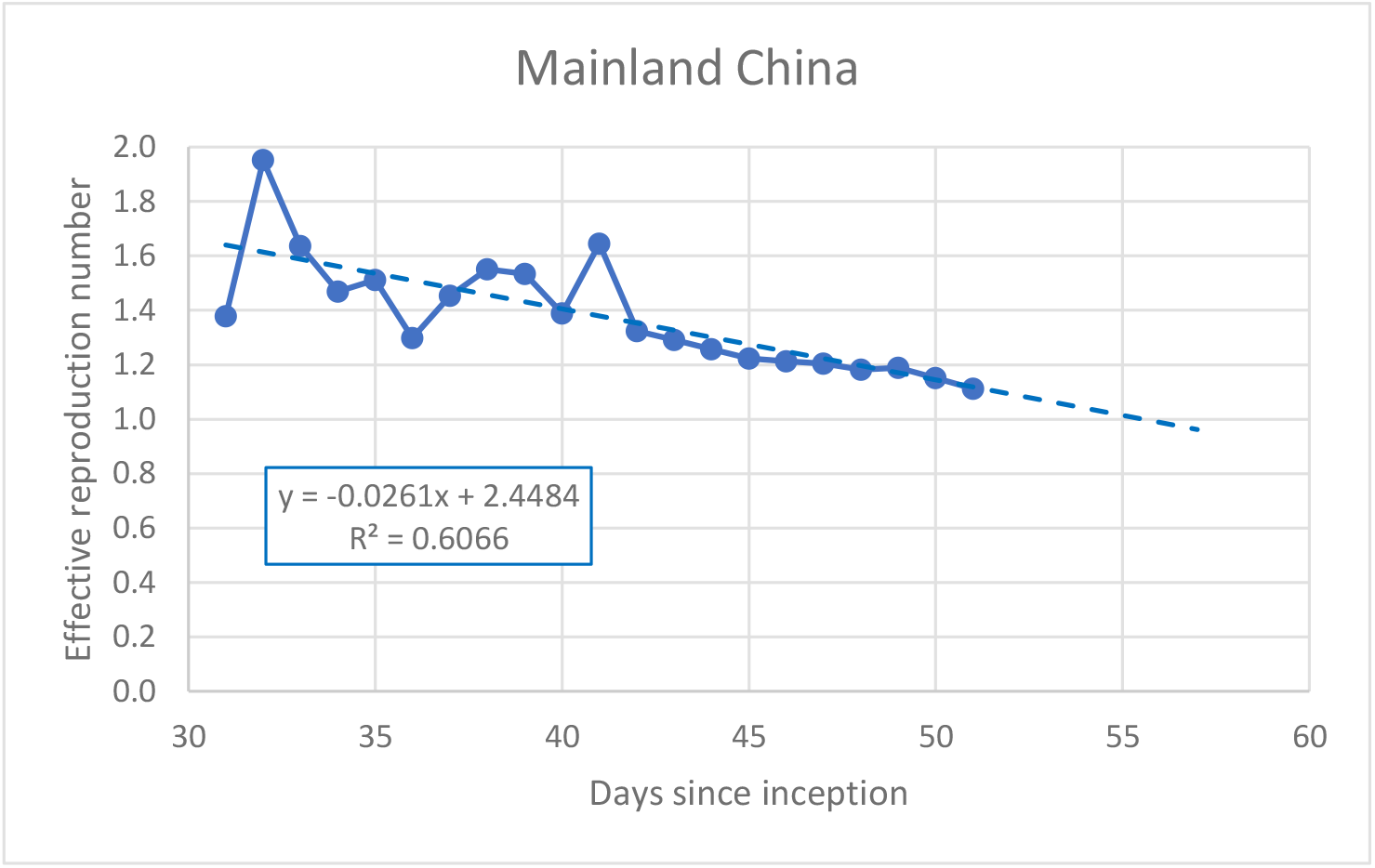
Linearization of the effective reproduction number with time

This linearization provides yet another method for the use of the logistic model. We use linear regression as shown in Figure *3* to calculate the effective reproduction numbers for a series of days *R*_*e*_^*\**^ *(t)* with the ^*^ superscript denoting that the values are calculated from the regression. Using xsEqs. (3) and (4) the predicted values of *P(t)* can be calculated regressively as

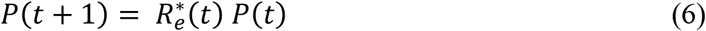

### Forecast and Discussion

Based on the linearization of Eq. (5), we obtained the parameters of the logistic model for three periods as shown in Table 2. The resulting prediction of future infection cases are shown in Figure *4*

**Figure 4.**
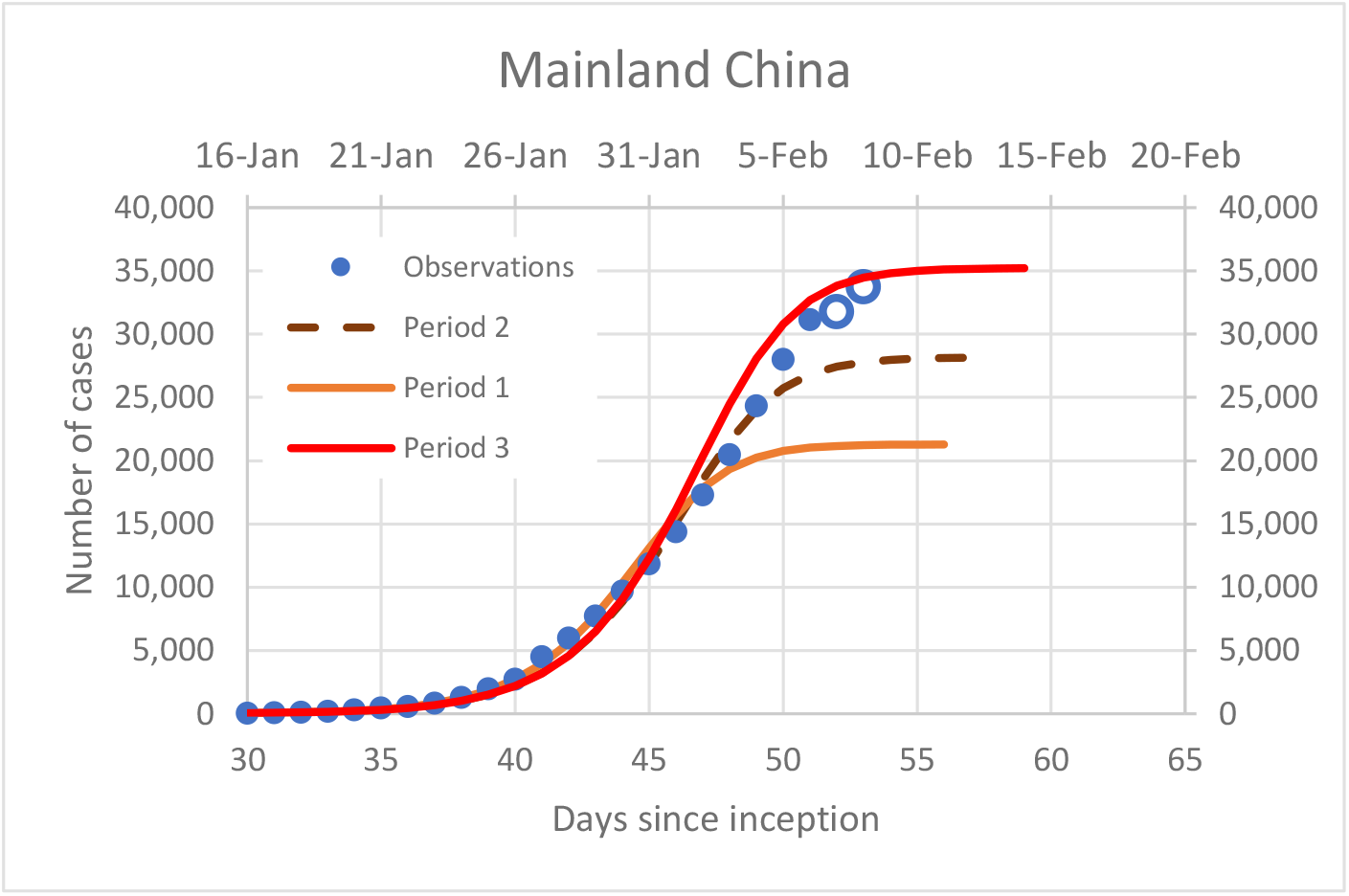
Recorded infection cases and model predictions for three periods analyzed. Data up to day 51 = February 6 were used for model fitting. Open circle – newest data point not included in the model fitting

**Table 2.** Consecutive model refinements

| Period End | | $R_0^*$ | $K$ | Maximum cases | Peak time (day) |
| --- | --- | --- | --- | --- | --- |
| Day | Date |  |  |  |  |
| 45 | January 31 | 1.52 | $6.2 \cdot 10^4$ | $3.52 \cdot 10^4$ | 56 |
| 48 | February 3 | 1.48 | $8.6 \cdot 10^4$ | $2.81 \cdot 10^4$ | 57 |
| 51 | February 6 | 1.48 | $1.1 \cdot 10^5$ | $3.52 \cdot 10^4$ | 60 |

The model predicted further increases of the infected cases but slower than the initial exponential growth. As expected, new data resulted in further model refinement primarily in the values of *K* and the resulting maximum cases.

Following a parallel approach and using Eq. (6) we obtained yet another set of predictions shown in together with those from Figure *4*. This approach results in a dramatically higher predictions of maximum case (at about 65,000) but remarkably shows a very close time for the peak (Figure *5*).

**Figure 5.**
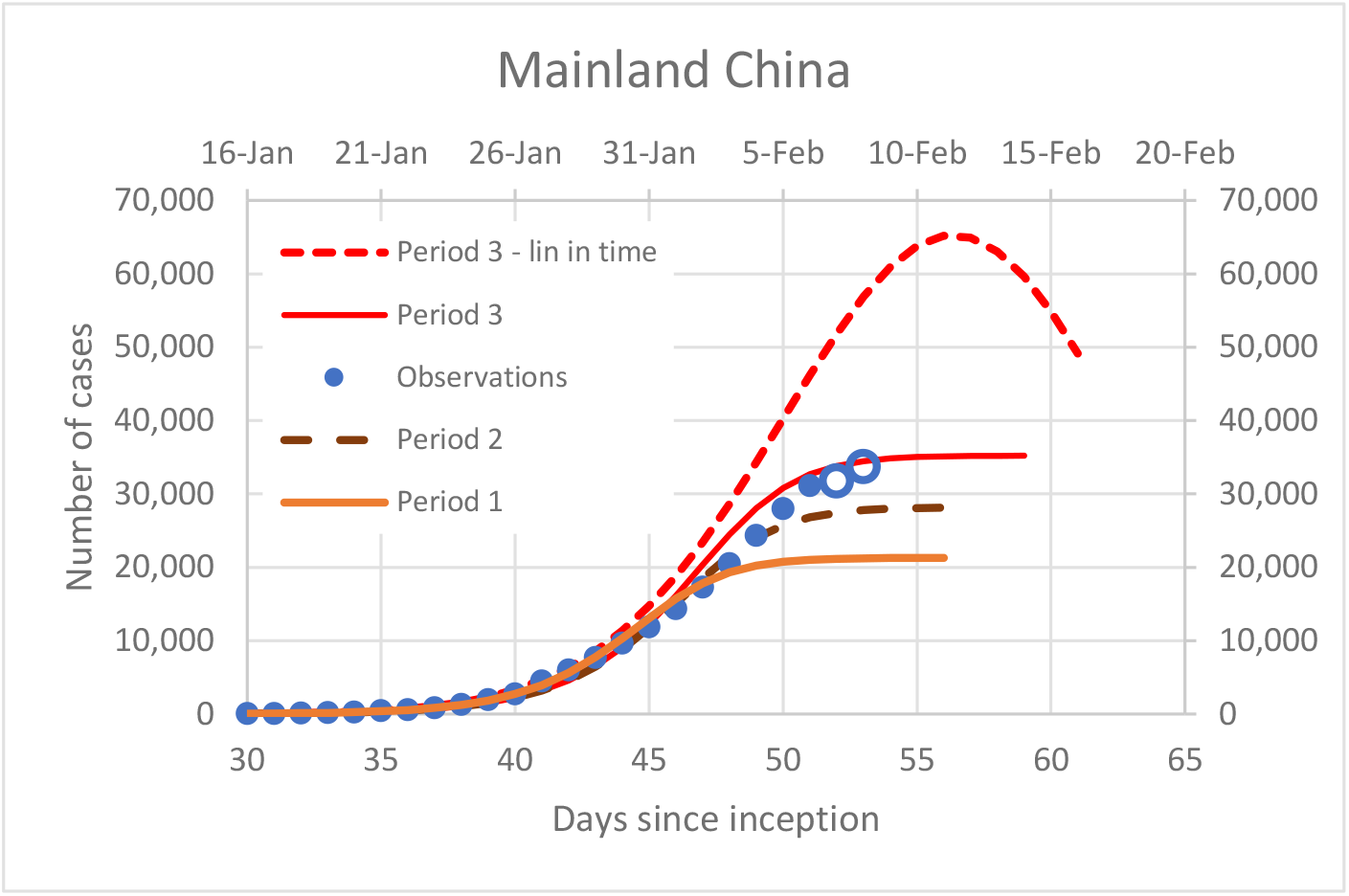
Predictions of model using linearization in time (Eq. 6) together with those of Figure 4. Open circle – newest data point not included in the model fitting

Despite differences among different estimate, the **critical feature** of the model predictions is the **stabilization** of the total number of cases in the next several days (by mid-February) and not a dramatic exponential growth. This prediction is made purely based on the described analysis and may or may not happen in the future. One significant factor that could invalidate the predictions is a possibility of secondary or parallel outbreaks perhaps with different etiology as previously suggested for the original Wuhan outbreak (P. Wu et al., 2020)

## Data Availability

not applicable - all data in public domain

## Declarations

### Ethics approval and consent to participate

The ethical approval or individual consent was not applicable.

### Availability of data and materials

All data and materials used in this work were publicly available.

### Consent for publication

Not applicable.

### Disclaimer

The funding agencies had no role in the design and conduct of the study; collection, management, analysis, and interpretation of the data; preparation, review, or approval of the manuscript; or decision to submit the manuscript for publication.

